# Device-Measured Physical Activity in 3,511 Individuals with Knee or Hip Arthroplasty

**DOI:** 10.1101/2023.05.26.23290524

**Authors:** Scott R. Small, Sara Khalid, Andrew J. Price, Aiden Doherty

## Abstract

**Background:** Physical activity is a key contributor to overall health, with low activity levels associated with a higher risk of all-cause mortality. Hip and knee arthroplasty aims to reduce joint pain, increasing functional mobility and the ability to be more physically active. However, the degree to which arthroplasty is associated with higher physical activity is unclear. The current study sought to assess the association of hip and knee arthroplasty with objectively measured physical activity.

**Methods:** This cross-sectional study analysed seven days of wrist-worn accelerometer data collected in 2013-2016 from UK Biobank participants (aged 43-78), with a median 6.8 years of follow-up. From a cohort of 94,794 participants with valid accelerometer wear time and complete self-reported data, electronic health records were used to identify 3,511 participants having undergone primary or revision hip or knee arthroplasty and 68,450 non-arthritic controls. Multivariable linear regression was performed to assess activity in terms of steps, cadence, acceleration, and behavioural classification between end-stage arthritic and >12 month postoperative cohorts, controlling for demographic and behavioural confounders.

**Findings:** Any combination of one or more hip or knee arthroplasties was associated with participants taking 694 more daily steps [95% CI: 346, 1,041] (p<0.001) than participants with end-stage osteoarthritis, though these participants took 542 fewer daily steps [95% CI: 403, 680] (p<0.001) than non-arthritic controls. Unilateral primary hip and knee arthroplasty were associated with 858 [95% CI: 255, 1,461] (p=0.005) and 879 [95% CI: 209, 1,549] (p=0.010) more steps than end-stage osteoarthritic participants, respectively. Postoperative hip arthroplasty participants demonstrated levels of moderate-to-vigorous physical activity and cadence equivalent to non-arthritic controls.

**Interpretation:** Hip and knee arthroplasty are associated with higher levels of physical activity compared to participants with end-stage arthritis. Hip arthroplasty patients, in particular, approach equivalence with non-arthritic peers in peak cadence and time spent in moderate-to-vigorous activity. Accelerometer-based activity monitoring of arthroplasty patients may provide an objective means for the assessment of postoperative recovery and interventional effectiveness.

## Introduction

Physical activity and mobility are key areas of focus for rehabilitation and perioperative care to improve joint function and patient quality of life following hip and knee arthroplasty as treatment for end-stage osteoarthritis. Historically, physical activity in orthopaedic populations has been assessed primarily through the use of self-reported physical activity questionnaires^1^. Self-reported physical activity questionnaires suffer from significant recall bias and have been preferentially replaced with wearable-measured physical activity assessment in large-scale epidemiological studies^2, 3^. However, recent investigations of wearable-measured physical activity in lower limb arthroplasty clinical cohorts are predominantly composed of small (n<100), select, or nominally matched cohorts, leading to conflicting evidence regarding postoperative levels of physical activity following joint replacement surgery^1, 4, 5^.

While knee and hip arthroplasty are well-established surgical interventions ^6, 7^, key questions around physical activity in the clinical population remain unresolved, namely: 1) What is the impact of end-stage knee or hip osteoarthritis on physical activity compared to a peer non-arthritic population? 2) Does surgical intervention lead to elevated physical activity levels after full recovery? 3) Following recovery, do arthroplasty patients reach expected activity levels equivalent to their non-arthritic peers?

We therefore aimed to quantify differences in device-measured physical activity between non-arthritic controls, participants with end-stage hip or knee osteoarthritis, and participants having undergone hip or knee arthroplasty. We additionally aimed to address historical challenges of small sample size, selection bias, and poorly controlled confounders, which hinder the generalisability of many studies in the current orthopaedic literature.

## Methods

### Activity Measurement

Between 2013 and 2016, over 100,000 UK Biobank participants agreed to wear an Axivity AX3 accelerometer on their dominant wrist, recording at a sampling rate of 100 Hz and resolution of ± 8 *g* for 24 hours a day for 7 days. Acceleration metrics and machine learning-derived behavioural classification were retrieved from accelerometer-derived variables from the UK Biobank dataset^2, 8^. The metric of overall acceleration is generated using accelerometer data processed through a previously described series of resampling, calibration, and lowpass filtering^2^. A validated random forest and hidden Markov behaviour model was used to allocate time use into sleep, sedentary, light activity, and moderate-to-vigorous physical activity (MVPA) classifications^8^. Raw accelerometer data for each participant was additionally accessed for this study to calculate median daily step count and peak cadence using a hybrid self-supervised machine learning step detection algorithm that has previously demonstrated high accuracy in free-living validation^9^. For each participant, valid wear time was considered a contribution of at least 72 hours of accelerometer wear, in addition to a contribution of at least one hour of wear during each one-hour period over the 24-hour clock^2^. Participants with an improbable average acceleration of over 100 m*g* were excluded from the accelerometry cohort.

### Health Data Linkage

National Health Service Hospital Episode Statistics are linked with the UK Biobank, containing data from 1997 to 2021 for participants in England, 1998 to 2016 in Wales, and 1981 to 2021 in Scotland^10^. Operative codes are listed in the OPCS-4 coding classification system, while ICD-9 and ICD-10 codes are used for in-hospital admission episode diagnoses. A modified Charlson Comorbidity Index, ranging from 0-50, was used to quantify the multimorbidity status of all participants^11, 12^. Charlson Comorbidity Index was calculated based on ICD-10 data linkage for each participant within a 5-year lookback window from the start date of the participant’s accelerometer wear period.

### Cohort Generation

A flow diagram for study cohort generation is presented in Figure 1. OPCS-4 code definitions from the National Joint Registry^13^ were used to identify participants with any documented history of primary or revision knee or hip arthroplasty. Participants who wore an accelerometer less than 12 months after their first hip or knee arthroplasty were considered still in transient early recovery and were not included in this analysis. Additionally, participants who received their first hip or knee arthroplasty more than 12 months into the future following accelerometer wear were also removed.

**Figure 1:**
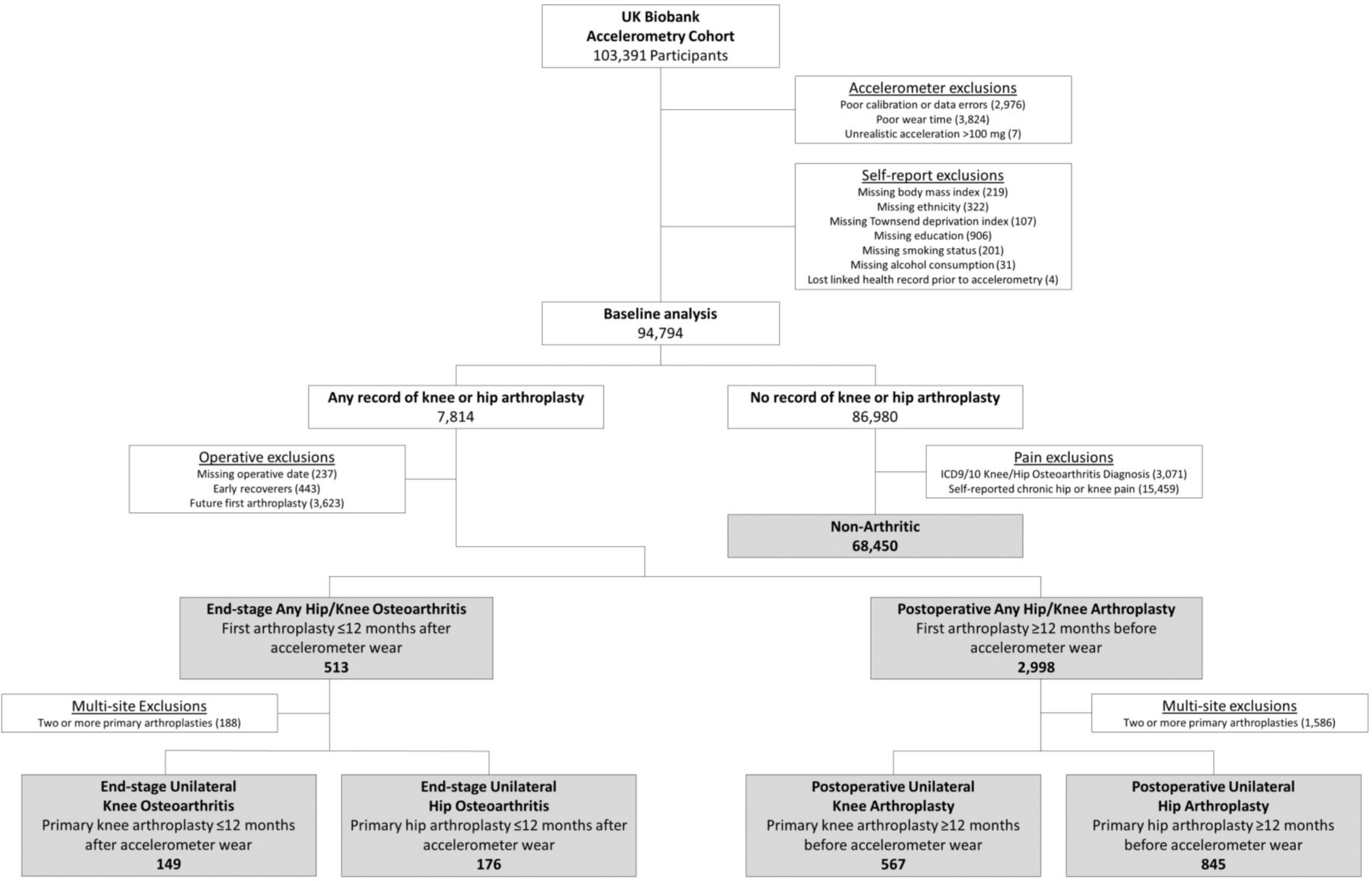
Participant flow chart. Non-arthritic and clinical participant subgroups based on operative status and timing relative to accelerometer wear date. Excluded early recoverers are considered participants who wore an accelerometer less than 12 months after their first primary hip or knee arthroplasty. Participants were also excluded if their first primary hip or knee arthroplasty was conducted more than 12 months after accelerometer wear.

To assess physical activity associated with any combination of primary or revision knee and hip arthroplasty, two initial study groups were created. The first group consisted of end-stage osteoarthritic participants who underwent their first primary knee or hip arthroplasty less than 12 months after accelerometer wear. The second group consisted of participants with any number of hip or knee arthroplasties, having had their first primary arthroplasty at least 12 months before accelerometer wear. For a second analysis in more clearly defined cohorts, participants with only a single, unilateral primary knee or hip arthroplasty at any point in their medical records were selected from the initial cohorts. Participants who had one or more subsequent revisions of the single primary arthroplasty were also included. Once arthroplasty patients were identified, cohorts were delineated into four subgroups: end-stage unilateral knee osteoarthritis with no arthroplasty at the time of accelerometer wear, end-stage unilateral hip osteoarthritis with no arthroplasty at the time of accelerometer wear, postoperative with a single-site primary or revised knee arthroplasty at the time of accelerometer wear, and postoperative with a single-site primary or revised hip arthroplasty at the time of accelerometer wear.

A reference cohort of non-arthritic controls was selected for comparison with knee and hip arthroplasty cohorts. The non-arthritic reference cohort was defined as any UK Biobank participant with valid accelerometer wear time, no medical record of hip or knee arthroplasty up to the latest point of health record update, no hip or knee osteoarthritis diagnosis, and no self-report of chronic (3+ months) of hip pain or knee pain at any follow-up appointment or interaction with the UK Biobank. All combinations of OPCS and ICD codes used for cohort generation are listed in the supplemental materials.

### Statistical Analysis

Multivariable linear regression was performed to compare physical activity between selected clinical cohorts. Confounders for adjustment in the regression model were selected from UK Biobank variables having a significant correlation with physical activity in other epidemiological studies, including age at the time of wear, sex, ethnic background, education level, smoking status, alcohol consumption status, Townsend Deprivation Index, wear season, and Charlson Comorbidity Index^2, 8, 14^. Due to low overall missingness within the UK Biobank, a complete-case analysis was performed; participants with missing data in any of the confounding exposure variables were removed from the analysis. Primary outcome metrics of interest were step count, peak cadence, overall acceleration, and machine learning classified moderate-to-vigorous physical activity. Additional measures of machine learning classified light activity, sedentary activity, and sleep were quantified as a secondary analysis. Separate regression models for each physical activity outcome measure were generated. All statistical analysis was performed using R (v4.1.0) on the Biomedical Research Cluster at the University of Oxford Big Data Institute.

## Results

Baseline demographics of the UK Biobank clinical cohorts with valid accelerometer wear and self-reported demographic data are presented in Table 1. A total of 7,814 participants were identified as having any record of knee or hip arthroplasty, with 3,511 having their first arthroplasty at least 12 months before or within 12 months after accelerometer wear. Of these, 149 participants contributed valid accelerometer wear less than 12 months before the primary knee arthroplasty, while 567 participants wore the accelerometer 12 or more months after the primary knee arthroplasty. A total of 176 participants wore an accelerometer less than 12 months before a single unilateral hip arthroplasty and 845 wore an accelerometer more than 12 months after a single hip arthroplasty. To serve as a reference baseline, 68,450 participants were identified as having had no hip or knee arthroplasty, lower limb osteoarthritis, or chronic hip or knee pain. Consistent with the broader UK Biobank cohort, all clinical subgroups consisted of primarily white ethnicity with approximately 50% of participants coming from the least deprived quintile of the broader population of the United Kingdom.

**Table 1:** Study cohort demographics

| Characteristic | Any Hip or Knee Arthroplasty<sup>+</sup> |  | Single Knee Arthroplasty<sup>*</sup> |  | Single Hip Arthroplasty<sup>*</sup> |  | Non-Arthritic Cohort |
| --- | --- | --- | --- | --- | --- | --- | --- |
|  | End-Stage Arthritis<sup>#</sup> | 12+ Months Postop<sup>‡</sup> | End-Stage Arthritis<sup>#</sup> | 12+ Months Postop<sup>‡</sup> | End-Stage Arthritis<sup>#</sup> | 12+ Months Postop<sup>‡</sup> |  |
| Overall, N (%) | 513 (100) | 2,998 (100) | 149 (100) | 567 (100) | 176 (100) | 845 (100) | 68,450 (100) |
| Sex, N (%) |  |  |  |  |  |  |  |
| Female | 301 (59) | 1,720 (57) | 79 (53) | 304 (54) | 103 (59) | 481 (57) | 38,203 (56) |
| Male | 212 (41) | 1,278 (43) | 70 (47) | 263 (46) | 73 (41) | 364 (43) | 30,247 (44) |
| Age, Median [IQR] | 67.9 [64.0 – 71.4] | 68.8 [64.9 – 72.1] | 68.3 [65.0–71.6] | 69.3 [65.7–72.3] | 67.6 [62.7–71.5] | 68.3 [64.2–71.8] | 62.6 [55.4–68.1] |
| Ethnicity, N (%) |  |  |  |  |  |  |  |
| White | 508 (99) | 2,954 (99) | 146 (98) | 555 (98) | 176 (100) | 836 (99) | 66,276 (97) |
| Non-white | 5 (1) | 44 (1) | 3 (2) | 12 (2) | 0 (0) | 9 (1) | 2,174 (3) |
| Body Mass Index, Median [IQR] | 28.2 [25.2 – 31.7] | 28.3 [25.5 – 31.7] | 28.3 [26.0–31.8] | 29.2 [26.4–32.9] | 26.8 [24.7–29.6] | 26.7 [24.2–29.8] | 25.6 [23.3–28.4] |
| CCI, Median [IQR] | 0 [0–0] | 0 [0–4] | 0 [0–4] | 0 [0–4] | 0 [0–0] | 0 [0–0] | 0 [0–0] |
| Wear Season, N (%) |  |  |  |  |  |  |  |
| Spring | 121 (24) | 653 (22) | 37 (25) | 113 (20) | 42 (24) | 175 (21) | 15,819 (23) |
| Summer | 152 (30) | 823 (27) | 40 (27) | 159 (28) | 53 (30) | 232 (27) | 18,083 (26) |
| Autumn | 136 (27) | 898 (30) | 39 (26) | 177 (31) | 48 (27) | 263 (31) | 20,195 (30) |
| Winter | 104 (20) | 624 (21) | 33 (22) | 118 (21) | 33 (19) | 175 (21) | 14,353 (21) |
| Education, N (%) |  |  |  |  |  |  |  |
| School Leaver | 152 (30) | 924 (31) | 39 (26) | 206 (36) | 57 (32) | 250 (30) | 14,823 (22) |
| Further Education | 170 (33) | 1,066 (36) | 58 (39) | 210 (37) | 39 (22) | 274 (32) | 22,292 (33) |
| Higher Education | 191 (37) | 1,008 (34) | 52 (35) | 151 (27) | 80 (45) | 321 (38) | 31,335 (46) |
| Smoking Status, N (%) |  |  |  |  |  |  |  |
| Never | 242 (47) | 1,496 (50) | 78 (52) | 267 (47) | 86 (49) | 455 (54) | 40,354 (59) |
| Former | 236 (46) | 1,335 (45) | 60 (40) | 271 (48) | 81 (46) | 337 (40) | 23,463 (34) |
| Current | 35 (7) | 167 (6) | 11 (7) | 29 (5) | 9 (5) | 53 (6) | 4,633 (7) |
| Alcohol Consumption, N (%) |  |  |  |  |  |  |  |
| Never | 29 (6) | 205 (7) | 8 (5) | 40 (7) | 9 (5) | 46 (5) | 3,748 (5) |
| < 3 Days Per Week | 233 (45) | 1,351 (45) | 69 (46) | 270 (48) | 74 (42) | 355 (42) | 30,803 (45) |
| 3+ Days Per Week | 251 (49) | 1,442 (48) | 72 (48) | 257 (45) | 93 (53) | 444 (53) | 33,899 (50) |
| Townsend Deprivation<sup>34</sup>, N (%) |  |  |  |  |  |  |  |
| Least Deprived | 251 (49) | 1,551 (52) | 71 (48) | 280 (49) | 83 (47) | 458 (54) | 34,638 (51) |
| 2<sup>nd</sup> Quintile | 136 (27) | 673 (22) | 43 (29) | 126 (22) | 47 (27) | 182 (22) | 15,385 (22) |
| 3<sup>rd</sup> Quintile | 64 (12) | 400 (13) | 15 (10) | 89 (16) | 23 (13) | 110 (13) | 9,709 (14) |
| 4<sup>th</sup> Quintile | 46 (9) | 278 (9) | 14 (9) | 59 (10) | 15 (9) | 66 (8) | 6,490 (9) |
| Most Deprived | 16 (3) | 96 (3) | 6 (4) | 13 (2) | 8 (5) | 29 (3) | 2,228 (3) |

Percentages may not add up to 100% due to rounding
CCI: Charlson Comorbidity Index
+ Includes all participants with any combination of primary or revision knee or hip arthroplasties, excluding those who had their first arthroplasty less than 12 months before accelerometer wear (early recoverers), or more than 12 months after accelerometer wear (future first arthroplasty).
\*Single Hip and Single Knee Arthroplasty Cohorts are subsets of the Any Hip or Knee Arthroplasties Cohort as defined in Figure 1
<sup>#</sup> End-stage arthritis is considered having their first arthroplasty within 12 months after accelerometer wear
<sup>‡</sup> Accelerometer wear more than 12 months after the participant's first primary knee or hip arthroplasty

### Physical Activity and Mobility Associated with Any Hip or Knee Arthroplasty

Adjusted estimated marginal means of step count, peak cadence, overall acceleration, and machine learning-derived time spent in MVPA, derived from linear regression models, are presented in Figure 2. Unadjusted activity levels for each cohort and full regression results are presented in the supplement (Tables S3-S5). Participants with any end-stage hip or knee arthroplasty took 1,236 fewer daily steps [95% CI: 912, 1,559] (p<0.001), had a peak cadence of 2.9 fewer steps per minute [95% CI: 2.0, 3.7] (p<0.001), and participated in 5.8 fewer minutes per day of MVPA [95% CI:2.9, 8.6] (p<0.001) compared to non-arthritic controls. Postoperative participants took 542 [95% CI: 403, 680] (p<0.001) more daily step counts and 1.8 [95% CI: 1.5, 2.2] (p=0.019) more steps per minute peak cadence compared with end-stage osteoarthritic participants; however, participants who had undergone any combination of hip or knee arthroplasty were less active than non-arthritic peers in terms of daily step count, peak cadence, and time spent in MVPA. No statistically significant difference in overall acceleration was observed in any comparison.

**Figure 2:**
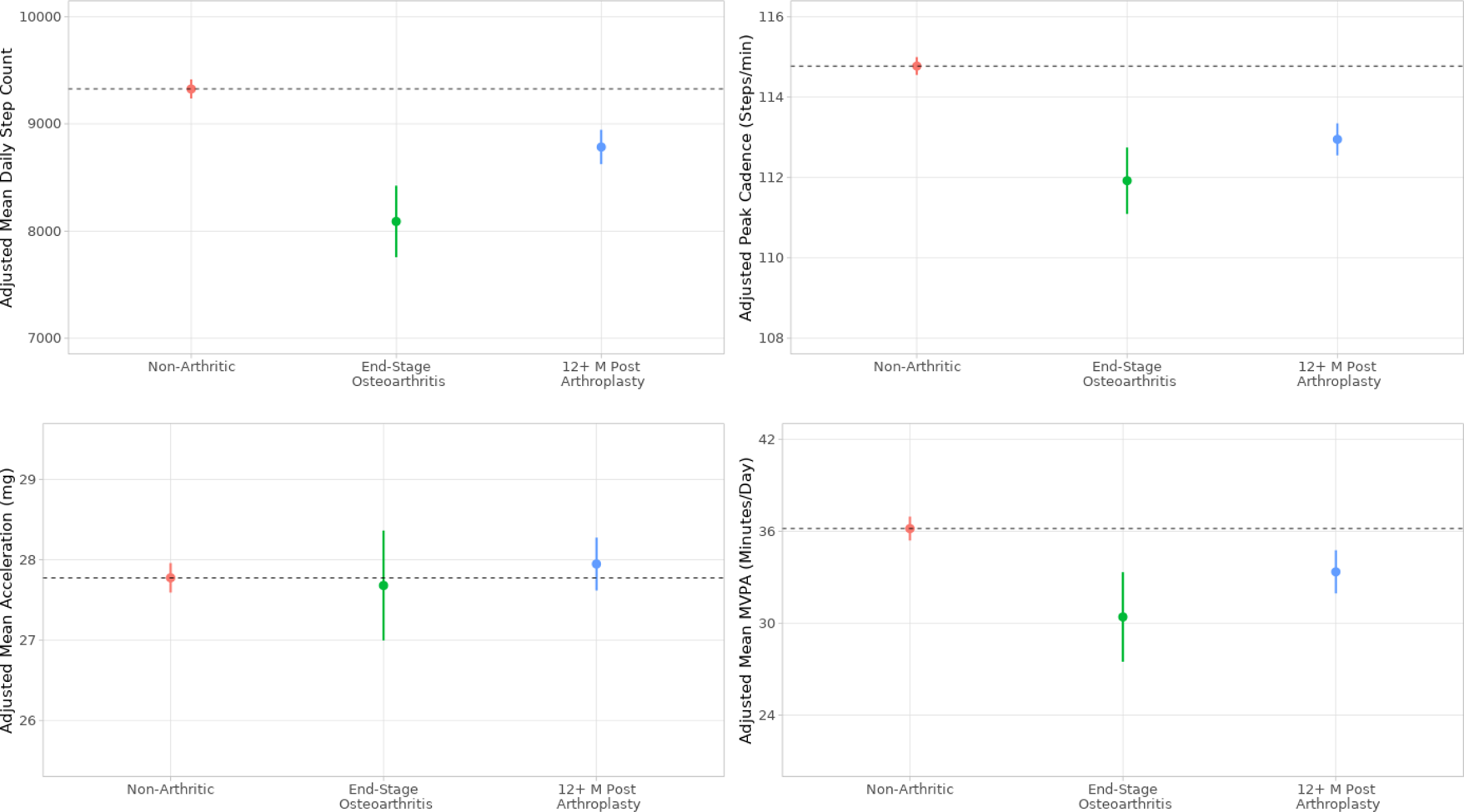
Estimated marginal means and 95% confidence intervals of activity and mobility metrics in participants who are non-arthritic, end-stage osteoarthritic, and more than 12 months postoperative from first primary knee or hip arthroplasty. Metrics are adjusted for age, sex, body mass index, season, Charlson Comorbidity Index, education, ethnicity, alcohol consumption, and smoking status.

### Physical Activity and Mobility Associated with a Single, Unilateral Hip or Knee Arthroplasty

Adjusted estimated marginal means of linear regression-derived step count, peak cadence, overall acceleration, and machine learning-derived time spent in MVPA are presented in Figure 3. Full results from the linear regression models are presented in Supplemental Tables S6 and S7. Following adjustment for covariates, end-stage unilateral knee osteoarthritis was associated with lower physical activity when compared to the non-arthritic reference cohort, taking 1,387 fewer daily steps [95% CI: 789, 1,984] (p<0.001), a peak cadence of 4.1 [95% CI: 2.7, 5.6] (p<0.001) fewer steps per minute, and 6.8 [95% CI: 1.5, 12.0] (p=0.012) fewer daily minutes spent in MVPA. End-stage unilateral hip osteoarthritis was associated with taking 1,134 [584, 1,683] (p<0.001) fewer daily steps than controls, but participants had otherwise statistically similar physical activity to their non-arthritic peers. Unilateral primary hip and knee arthroplasty were associated with 858 [95% CI: 255, 1,461] (p=0.005) and 879 [95% CI: 209, 1,549] (p=0.010) more steps than end-stage osteoarthritic participants, respectively. However, postoperative unilateral knee arthroplasty was associated with 508 [95% CI: 199, 816] (p=0.001) fewer daily steps, a cadence of 2.8 [95% CI: 2.0, 3.6] (p<0.001) fewer steps per minute, and 2.9 [0.2, 5.6] (p=0.037) fewer daily minutes of MVPA compared to controls. Postoperative unilateral hip arthroplasty was associated with a modest 276 [95% CI: 23, 529] (p=0.033) fewer daily steps than controls, but these participants were otherwise equivalent to non-arthritic peers in terms of activity and mobility. No difference in physical activity was observed between any cohorts in terms of overall daily light activity, sedentary behaviour, or sleep time.

**Figure 3:**
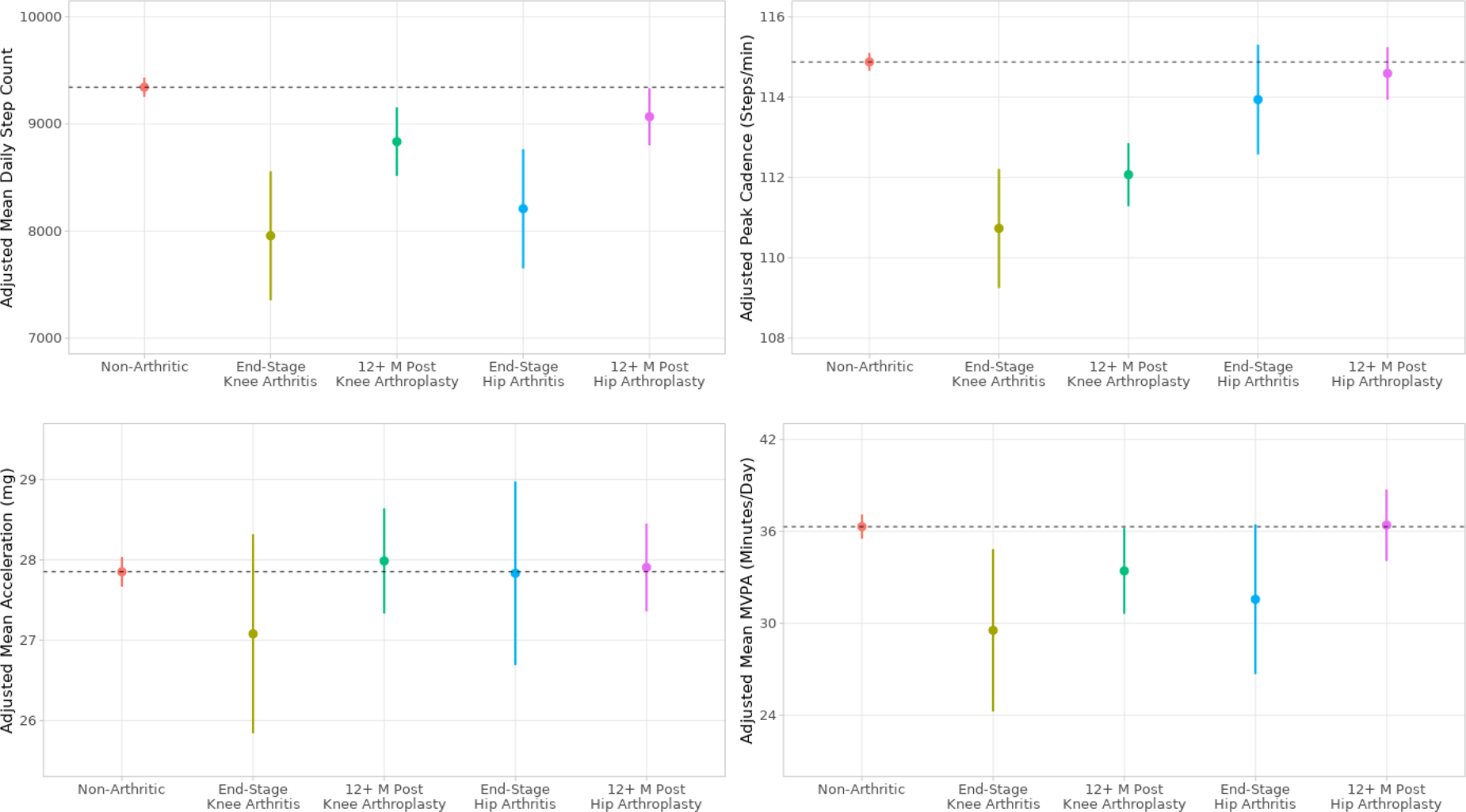
Estimated marginal means and 95% confidence intervals of activity and mobility metrics in participants who are non-arthritic, have end-stage knee osteoarthritis, are more than 12 months postoperative from unilateral primary knee arthroplasty, have end-stage hip osteoarthritis, or are more than 12 months postoperative from unilateral primary hip arthroplasty. Metrics are adjusted for age, sex, body mass index, season, Charlson Comorbidity Index, education, ethnicity, alcohol consumption, and smoking status.

## Discussion

Our cross-sectional study indicated significantly higher activity in terms of daily step count following hip and knee arthroplasty when compared to participants with end-stage hip osteoarthritis. As a result, participants who have undergone surgical intervention demonstrate only modestly lower or equivalent physical activity compared to their non-arthritic peers across metrics, with the most room for improvement seen in postoperative knee arthroplasty cohorts in terms of daily step count, peak cadence, and time spent in moderate-to-vigorous physical activity.

The first objective of this study was to assess the effect of end-stage knee and hip arthritis on physical activity compared to a non-arthritic peer group. We found that participants with any end-stage hip or knee osteoarthritis took 13% fewer steps per day, had a 3% slower peak cadence, and spent 16% fewer minutes per day in moderate-to-vigorous physical activity compared to non-arthritic peers. When assessing activity in participants with single-site arthritis, both knee and hip arthritis were associated with lower daily steps. Only end-stage knee arthritis was associated with lower peak cadence and time spent in moderate-to-vigorous physical activity compared to non-arthritic controls. Prior, smaller studies of objectively measured physical activity in end-stage osteoarthritic patients show similar patterns, yet greater differences, with end-stage hip or knee osteoarthritis associated with 20-35% fewer steps and active minutes than health controls^15–17^.

When comparing postoperative unilateral knee and hip arthroplasty cohorts to end-stage arthritic cohorts, our results indicate 11% and 10% higher daily step count in the postoperative unilateral knee and hip arthroplasty cohorts, respectively. Prior evidence as to whether physical activity increases for the average patient following surgery has been mixed, as most historical investigations of physical activity in patients with hip and knee arthroplasty have been conducted using self-reported questionnaires limited by well-documented methodological biases.^1, 3^ In these cases, many patients report increased physical activity and functional improvement on patient reported outcome measures; however, the majority of studies using objectively measured activity describe only minimal gains in postoperative activity and step count in small patient populations^1, 18, 19^. In a longitudinal study using a consumer-grade activity tracker, Christensen et al.^5^ reported a return to preoperative step count within one month following unilateral knee arthroplasty in 1,005 total knee arthroplasty patients, culminating in approximately 40% more daily steps at one-year follow-up. This increase is substantially higher than studies of previous cohorts, and may be a result of selection bias by including only participants who owned Apple Watch-compatible iPhones at targeted surgical centres. A more moderate difference in activity levels between postoperative and end-stage arthritic cohorts was observed in the current study, which may result, in part, from the UK Biobank drawing health records from surgical interventions performed across the breadth of public healthcare facilities.

An important target outcome for successful lower limb arthroplasty is the restoration of joint mobility, enabling the potential of increased physical activity and improved overall patient health compared to before surgical intervention. Physical activity is well correlated with health outcomes and an increase of 1,000 steps per day has been attributed to a lower risk of cardiovascular disease and all-cause mortality^20^. Compared to their non-arthritic peers, postoperative unilateral knee arthroplasty was associated with taking 5% fewer daily steps, 2.8 fewer steps per minute slower cadence, and 8% less time in MVPA. Conversely, postoperative unilateral hip arthroplasty was associated with only 3% fewer daily steps and equivalent peak cadence and levels of MVPA compared to non-arthritic controls. Other comparisons have been drawn in terms of physical activity levels between postoperative hip or knee arthroplasty patients and non-arthritic controls, reporting significantly lower activity in these patients compared to their peers^21–26^. Walker et al.^24^ and Kersten et al.^26^ reported that activity in postoperative knee patients may still be up to 20% less than in healthy controls. Whereas most prior studies of physical activity in orthopaedic populations to healthy controls have generally matched only on age and sex^1, 23^, the current analysis adjusts for other confounders of activity, including body mass index, education, alcohol intake, smoking status, and medical comorbidities^8^.

In this study, daily step count, time spent in MVPA and peak cadence metrics demonstrated sensitivity to lower limb osteoarthritis and surgical intervention, yet no differences in physical activity were observed between any cohorts when comparing overall acceleration magnitude. While overall acceleration is a validated surrogate for general activity level^27^, the inclusion of mobility-centric metrics like step count and peak cadence may enable more nuanced analysis in certain populations. This may imply that orthopaedic surgical intervention affects overall mobility but does not greatly affect behavioural patterns of sleep, sedentary time, and light activity. In future studies, the inclusion of step metrics and machine learning classification of behaviours, such as time spent in MVPA, may elucidate insights into physical activity not distinguished by acceleration alone.

This study incorporates a highly accurate free-living validated step-counting algorithm designed from the same research-grade accelerometer as used in the UK Biobank; however, some caution may need to be taken when applying a step detection algorithm to orthopaedic clinical populations^28–30^, as up to 16% of arthroplasty patients have at least some dependence on walking aids^31^. Because assistance is generally minor^31–33^, usually consisting of a single cane^33^, the effect on step count accuracy will be small compared to more substantial gait aids^30^. Lower step count in participants with end-stage osteoarthritis may include some underestimation bias due to walking aid use. Even so, this increased postoperative step count retains and supports the overall conclusion of improved functional mobility (increased activity and/or reduced levels of aid) in postoperative arthroplasty patients. Of note, the UK Biobank health record linkage lacks data on privately accessed medical services, thus an unknown level of missing operative records from private healthcare facilities are unaccounted for, particularly within the non-arthritic control population. The use of additional ICD diagnostic codes alongside the conservative exclusion of any participants with self-reported knee or hip pain was incorporated to minimise the inclusion of hidden operative cases within the cohort. The current study is a cross-sectional analysis and does not enable patient-level assessment of physical activity before and after knee or hip arthroplasty. Through physical activity monitoring and digital health interventions, opportunities exist for further carefully designed longitudinal evaluation of postoperative recovery to increase activity levels, improve mobility and identify patient outliers in need of further intervention.

## Conclusion

This is the largest investigation of wearable-measured physical activity in hip or knee arthroplasty patients to date, with well-established health record linkage and detailed demographic data allowing for adjustment across multiple confounders of physical activity. The results highlight the significantly higher levels of activity in postoperative compared to end-stage osteoarthritic cohorts, with unilateral knee and hip arthroplasty patients taking approximately 850 steps per day more than respective end-stage arthritic peers. However, postoperative knee arthroplasty was associated with taking fewer steps per day than non-arthritic peers, providing motivation for further work in developing rehabilitation programs focused on improving physical activity, particularly in knee patients.

## Data Availability

All data used in this study was collected or derived from the UK Biobank. Our open-source accelerometer processing tool to calculate validated step count from raw accelerometry data is available for use at https://github.com/OxWearables/stepcount.

## Funding Acknowledgements

This research has been conducted using the UK Biobank Resource under Application Number 59070. This research was funded in whole or in part by the Wellcome Trust [223100/Z/21/Z]. For the purpose of open access, the author has applied a CC-BY public copyright licence to any author accepted manuscript version arising from this submission. AD and SS are supported by the Wellcome Trust. AD, SS, and SK are supported by HDR UK, an initiative funded by UK Research and Innovation, Department of Health and Social Care (England) and the devolved administrations. AD is further supported by Novo Nordisk, Swiss Re, and by the BHF Centre of Research Excellence (grant number RE/18/3/34214). SS is further supported by the University of Oxford Clarendon Fund. SK is further supported by Amgen and UCB BioPharma outside of the scope of this work. Computational aspects of this research were funded by the National Institute for Health Research (NIHR) Oxford Biomedical Research Centre (BRC) with additional support from Health Data Research (HDR) UK and the Wellcome Trust Core Award [grant number 203141/Z/16/Z]. The views expressed are those of the author(s) and not necessarily those of the NHS, the NIHR or the Department of Health.

