## supplemental materials for "Device-Measured Physical Activity in 3,511 Individuals with Knee or Hip Arthroplasty"

UK Biobank Fields ... ii
ICD-9 and ICD-10 Codes ... iii
Table S1: OPCS-4 Standalone Codes of Operation and Location ... iv
Table S2: OPCS-4 Combination Codes of Operation and Location ... v
Table S3: Unadjusted overall physical activity and mobility metrics by clinical cohort ... vi
Table S4: Linear regression adjusted differences between non-arthritic participants and participants with any combination of hip and/or knee arthroplasties in primary activity measures ... vii
Table S5: Linear regression adjusted differences in additional machine learning behavioural classifications between non-arthritic participants and participants with any combination of hip and/or knee arthroplasty ... viii
Table S6: Linear regression adjusted differences between non-arthritic participants and clinically-defined unilateral hip and knee participant cohorts in primary activity measures ... ix
Table S7: Linear regression adjusted differences in additional machine learning behavioural classifications between non-arthritic participants and clinically-defined unilateral hip and knee participant cohorts ... x

**UK Biobank Fields**

31 – Sex
34 – Year of Birth
52 – Month of Birth
189 – Townsend Deprivation Index at Recruitment
1558 – Alcohol Intake Frequency
3414 – Hip pain for 3+ months
3773 – Knee pain for 3+ months
6138 – Qualifications
20116 – Smoking Status
21000 – Ethnic Background
23101 – Body Mass Index
40046 – Sleep – Overall Average
40047 – Sedentary – Overall Average
40048 – Light – Overall average
40049 – Moderate-Vigorous – Overall Average
90001 - Accelerometer Data .cwa format (bulk)
90011 – End time of wear (accelerometer),
90015 – Data quality, good wear time (accelerometer)
90016 – Data quality good calibration (accelerometer)

**ICD9/10 Codes for Osteoarthritis (Hospital Episode Statistics)**

ICD-9 and ICD-10 diagnostic codes were used to identify diagnoses of hip or knee osteoarthritis at any time point. ICD-9 codes for hip and knee osteoarthritis included 71515 and 71516. ICD-10 codes for diagnosed knee arthritis included M170, M171, M172, M173, M174, M175, M179, and associated sublevels. ICD-10 codes for diagnosed hip arthritis included M160, M161, M162, M163, M164, M165, M166, M167, M169 and associated sublevels.

*Table S1: OPCS-4 Standalone Codes of Operation and Location*

| Primary Knee Arthroplasty | Revision Knee Arthroplasty | Primary Hip Arthroplasty | Revision Hip Arthroplasty |
|---|---|---|---|
| O181 | O180 | W371 | W370 |
| O188 | O182 | W378 | W372 |
| O189 | O183 | W379 | W373 |
| W401 | O184 | W381 | W374 |
| W408 | W400 | W388 | W380 |
| W409 | W402 | W389 | W382 |
| W411 | W403 | W391 | W383 |
| W418 | W404 | W398 | W384 |
| W421 | W410 | W399 | W392 |
| W428 | W412 | W931 | W393 |
| W429 | W413 | W938 | W395 |
| | W414 | W939 | W462 |
| | W420 | W941 | W472 |
| | W422 | W948 | W482 |
| | W432 | W949 | W932 |
| | W425 | W951 | W933 |
| | | W958 | W940 |
| | | W959 | W942 |
| | | | W943 |
| | | | W952 |
| | | | W953 |
| | | | W954 |

*Table S2: OPCS-4 Combination Codes of Operation and Location*

| Primary Knee Arthroplasty | Revision Knee Arthroplasty | Primary Hip Arthroplasty | Revision Hip Arthroplasty |
|---|---|---|---|
| W521 (with: Z846, Z765, Z845, Z844, Z774, or Z787) | W424 (with: Y032 or Y037) | W521, W531, W541, or W581 (with: Z843, Z761, or Z756) | W394 (with: Y032 or Y037) |
| | W522, W523, W532, W533, W542, W543, W544, W553, W564, W574, W582, W603, W613, W641, or W642 (with: Z846, Z765, Z845, Z844, Z774, or Z787) | | W522, W523, W533, W542, W543, W544, W572, W574, W582 (with: Z843, Z761, or Z756) |

*Table S3: Unadjusted overall physical activity and mobility metrics by clinical cohort*

| Activity Metric | Any Hip or Knee Arthroplasty+ | | Single Knee Arthroplasty* | | Single Hip Arthroplasty* | | Non-Arthritic Cohort |
|---|---|---|---|---|---|---|---|
| | End-Stage Osteoarthritis# | 12+ Months Postop‡ | End-Stage Osteoarthritis# | 12+ Months Postop‡ | End-Stage Osteoarthritis# | 12+ Months Postop‡ | |
| Step Count (steps/day) | 7,850 [5,540 – 10,004] | 8,210 [5,847 – 10,853] | 7,843 [5,781 – 9,438] | 8,250 [5,764 – 10,692] | 8,119 [5,860 – 10,333] | 9,027 [6,549 – 11,453] | 9,413 [7,160 – 12,037] |
| Peak Cadence (steps/min) | 112 [106 – 118] | 113 [106 – 119] | 110 [104 – 115] | 112 [106 – 117] | 114 [108 – 121] | 115 [109 – 121] | 117 [111 – 123] |
| Acceleration (m*g*) | 24.7 [20.4 – 29.5] | 24.6 [20.5 – 29.7] | 24.4 [19.2 – 27.7] | 24.0 [20.1 – 28.9] | 25.2 [21.5 – 30.2] | 25.4 [21.2 – 30.3] | 27.4 [22.8 – 32.9] |
| MVPA (min/day) | 20.0 [7.9 – 43.1] | 22.8 [8.8 – 44.7] | 23.6 [8.7 – 39.6] | 19.7 [8.0 – 40.1] | 19.8 [10.4 – 50.0] | 30.6 [12.3 – 53.0] | 34.9 [17.6 – 59.2] |
| Light Activity (hours/day) | 4.9 [4.0 – 6.0] | 4.9 [3.9 – 6.0] | 4.8 [3.7 – 5.9] | 4.9 [3.8 – 6.0] | 4.9 [4.1 – 5.9] | 4.9 [4.0 – 6.0] | 4.9 [3.9 – 6.1] |
| Sedentary Behaviour (hours/day) | 9.5 [8.4 – 10.7] | 9.5 [8.3 – 10.8] | 9.4 [8.4 – 10.8] | 9.5 [8.2 – 10.7] | 9.6 [8.6 – 10.4] | 9.4 [8.2 – 10.6] | 9.4 [8.2 – 10.6] |
| Sleep (hours/day) | 8.8 [8.1 – 9.6] | 8.8 [8.1 – 9.6] | 8.8 [8.1 – 9.7] | 8.9 [8.2 – 9.7] | 8.7 [8.1 – 9.5] | 8.8 [8.0 – 9.6] | 8.7 [8.1 – 9.5] |

*Activity metrics reported as unadjusted median [interquartile range]*

*+ Includes all participants with any combination of primary or revision knee or hip arthroplasties, excluding those who had their first arthroplasty less than 12 months before accelerometer wear (early recoverers), or more than 12 months after accelerometer wear (future first arthroplasty).*

**Single Hip and Single Knee Arthroplasty Cohorts are subsets of the Any Hip or Knee Arthroplasties Cohort as defined in Figure 1*

*# End-stage arthritis is considered having their first arthroplasty within 12 months after accelerometer wear*

*‡ Accelerometer wear more than 12 months after the participant's first primary knee or hip arthroplasty*

*Table S4: Linear regression adjusted differences between non-arthritic participants and participants with any combination of hip and/or knee arthroplasties in primary activity measures*

| Metric | Reference Cohort | End-Stage Knee and/or Hip Osteoarthritis | 12+ Months After First Knee or Hip Arthroplasty |
|---|---|---|---|
| Median Daily Step Count (steps/day) | Non-Arthritic | -1,236 [-1,559, -912] *(p<0.001)* | -542 [-680, -403] *(p<0.001)* |
| | End-Stage Knee and/or Hip Osteoarthritis | | 694 [346, 1,041] *(p<0.001)* |
| Peak Cadence (steps/min) | Non-Arthritic | -2.9 [-3.7, -2.0] *(p<0.001)* | -1.8 [-2.2, -1.5] *(p<0.001)* |
| | End-Stage Knee and/or Hip Osteoarthritis | | 1.0 [0.2, 1.9] *(p=0.019)* |
| Overall Acceleration (m*g*) | Non-Arthritic | -0.1 [-0.8, 0.6] *(p=0.777)* | 0.2 [-0.1, 0.5] *(p=0.236)* |
| | End-Stage Knee and/or Hip Osteoarthritis | | 0.3 [-0.4, 1.0] *(p=0.461)* |
| Moderate-to-Vigorous Activity (min/day) | Non-Arthritic | -5.8 [-8.6, -2.9] *(p<0.001)* | -2.8 [-4.0, -1.6] *(p<0.001)* |
| | End-Stage Knee and/or Hip Osteoarthritis | | 2.9 [-0.1, 6.0] *(p=0.058)* |

*Estimate [95% confidence interval] in activity metrics adjusted for age, sex, body mass index, season, Charlson comorbidity index, education, ethnicity, alcohol consumption, and smoking status. A negative difference indicates lower activity in the cohort of interest relative to the reference cohort.*

*Table S5: Linear regression adjusted differences in additional machine learning behavioural classifications between non-arthritic participants and participants with any combination of hip and/or knee arthroplasty*

| Metric | Reference Cohort | End-Stage Knee and/or Hip Osteoarthritis | 12+ Months After First Primary Arthroplasty |
|---|---|---|---|
| Light Activity (min/day) | Non-Arthritic | 5.6 [-2.5, 13.8] *(p=0.172)* | 5.6 [2.1, 9.0] *(p=0.002)* |
| | End-Stage Knee and/or Hip Osteoarthritis | | -0.1 [-8.8, 8.6] (p=0.984) |
| Sedentary Behaviour (min/day) | Non-Arthritic | 0.9 [-8.1, -9.9] *(p=0.844)* | 1.0 [-2.8, 4.9] *(p=0.595)* |
| | End-Stage Knee and/or Hip Osteoarthritis | | 0.1 [-9.5, 9.9] *(p=0.977)* |
| Sleep Time (min/day) | Non-Arthritic | -0.8 [-7.2, 5.6] *(p=0.810)* | -3.8 [-6.5, -1.0] *(p=0.007)* |
| | End-Stage Knee and/or Hip Osteoarthritis | | -3.0 [-9.9, 3.8] *(p=0.390)* |

*Estimate [95% confidence interval] in activity metrics adjusted for age, sex, body mass index, season, Charlson comorbidity index, education, ethnicity, alcohol consumption, and smoking status. A negative difference indicates lower activity in the cohort of interest relative to the reference cohort.*

*Table S6: Linear regression adjusted differences between non-arthritic participants and clinically-defined unilateral hip and knee participant cohorts in primary activity measures*

| Metric | Reference Cohort | End-Stage Knee Osteoarthritis | 12+ Months After Knee Arthroplasty | End-Stage Hip Osteoarthritis | 12+ Months After Hip Arthroplasty |
|---|---|---|---|---|---|
| Step Count (steps/day) | Non-Arthritic | -1,387 [-1,984, -789] *(p<0.001)* | -508 [-816, -199] *(p=0.001)* | -1,134 [-1,683, -584] *(p<0.001)* | -276 [-529, -23] *(p=0.033)* |
| | End-Stage Knee Osteoarthritis | | 879 [209, 1,549] *(p=0.010)* | 253 [-557, 1,063] *(p=0.541)* | 1,111 [464, 1,757] *(p<0.001)* |
| | 12+ Months Post Knee Arthroplasty | | | -626 [-1,254, 2] *(p=0.051)* | 232 [-164, 627] *(p=0.251)* |
| | End-Stage Hip Osteoarthritis | | | | 858 [255, 1,461] *(p=0.005)* |
| Peak Cadence (steps/min) | Non-Arthritic | -4.1 [-5.6, -2.7] *(p<0.001)* | -2.8 [-3.6, -2.0] *(p<0.001)* | -0.9 [-2.3, 0.4] *(p=0.174)* | -0.3 [-0.9, 0.3] *(p=0.366)* |
| | End-Stage Knee Osteoarthritis | | 1.3 [-0.3, 3.0] *(p=0.111)* | 3.2 [1.2, 5.2] *(p=0.002)* | 3.9 [2.3, 5.4] *(p<0.001)* |
| | 12+ Months Post Knee Arthroplasty | | | 1.9 [0.3, 3.4] *(p=0.018)* | 2.5 [1.5, 3.5] *(p<0.001)* |
| | End-Stage Hip Osteoarthritis | | | | 0.7 [-0.8, 2.1] *(p=0.389)* |
| Overall Acceleration (m*g*) | Non-Arthritic | -0.8 [-2.0, 0.5] *(p=0.217)* | 0.1 [-0.5, 0.8] *(p=0.677)* | -0.0 [-1.1, 1.1] *(p=0.975)* | 0.1 [-0.5, 0.6] *(p=0.838)* |
| | End-Stage Knee Osteoarthritis | | 0.9 [-0.5, 2.3] *(p=0.196)* | 0.8 [-0.9, 2.4] *(p=0.374)* | 0.8 [-0.5, 2.2] *(p=0.222)* |
| | 12+ Months Post Knee Arthroplasty | | | -0.2 [-1.4, 1.1] *(p=0.816)* | -0.1 [-0.9, 0.7] *(p=0.845)* |
| | End-Stage Hip Osteoarthritis | | | | 0.1 [-1.2, 1.3] *(p=0.909)* |
| Moderate-to-Vigorous Activity (min/day) | Non-Arthritic | -6.8 [-12.0, -1.5] *(p=0.012)* | -2.9 [-5.6, -0.2] *(p=0.037)* | -4.7 [-9.6, 0.1] *(p=0.055)* | 0.1 [-2.1, 2.3] *(p=0.932)* |
| | End-Stage Knee Osteoarthritis | | 3.9 [-2.0, 9.8] *(p=0.196)* | 2.0 [-5.1, 9.2] *(p=0.576)* | 6.9 [1.2, 12.6] *(p=0.018)* |
| | 12+ Months Post Knee Arthroplasty | | | -1.9 [-7.4, 3.7] *(p=0.511)* | 3.0 [-0.5, 6.5] *(p=0.092)* |
| | End-Stage Hip Osteoarthritis | | | | 4.8 [-0.5, 10.1] *(p=0.074)* |

*Difference [95% confidence interval] in activity metrics adjusted for age, sex, body mass index, season, Charlson comorbidity index, education, ethnicity, alcohol consumption, and smoking status. A negative difference indicates lower activity in the cohort of interest relative to the reference cohort.*

*Table S7: Linear regression adjusted differences in additional machine learning behavioural classifications between non-arthritic participants and clinically-defined unilateral hip and knee participant cohorts*

| Metric | Reference Cohort | End-Stage Knee Osteoarthritis | 12+ Months After Knee Arthroplasty | End-Stage Hip Osteoarthritis | 12+ Months After Hip Arthroplasty |
|---|---|---|---|---|---|
| Light Activity (min/day) | Non-Arthritic | -0.4 [-15.4, 14.6] (p=0.960) | 6.1 [-1.6, 13.9] (p=0.121) | 3.5 [-10.2, 17.3] (p=0.614) | -0.4 [-6.7, 6.0] (p=0.910) |
| | End-Stage Knee Osteoarthritis | | 6.5 [-10.3, 23.3] (p=0.447) | 3.9 [-16.4, 24.4] (p=0.705) | 0.0 [-16.2, 16.2] (p=0.998) |
| | 12+ Months Post Knee Arthroplasty | | | -2.6 [-18.3, 13.2] (p=0.747) | -6.5 [-16.4, 3.4] (p=0.199) |
| | End-Stage Hip Osteoarthritis | | | | -3.9 [-19.0, 11.2] (p=0.612) |
| Sedentary Behaviour (min/day) | Non-Arthritic | 4.2 [-12.4, 20.8] (p=0.620) | -4.6 [-13.2, 4.0] (p=0.293) | 3.3 [-12.0, 18.6] (p=0.674) | 3.4 [-3.6, 10.4] (p=0.343) |
| | End-Stage Knee Osteoarthritis | | -8.8 [-27.5, 9.8] (p=0.354) | -0.9 [-23.5, 21.6] (p=0.937) | -0.8 [-18.8, 17.2] (p=0.931) |
| | 12+ Months Post Knee Arthroplasty | | | 7.9 [-9.6, 25.4] (p=0.376) | 8.0 [-3.0, 19.0] (p=0.153) |
| | End-Stage Hip Osteoarthritis | | | | 0.1 [-16.7, 16.9] (p=0.989) |
| Sleep Time (min/day) | Non-Arthritic | 3.0 [-8.8, 14.7] (p=0.622) | 1.4 [-4.7, 7.4] (p=0.660) | -2.1 [-12.9, 8.7] (p=0.704) | -3.1 [-8.1, 1.8] (p=0.216) |
| | End-Stage Knee Osteoarthritis | | -1.6 [-14.8, 11.6] (p=0.813) | -5.0 [-21.0, 10.9] (p=0.535) | -6.1 [-18.8, 6.6] (p=0.348) |
| | 12+ Months Post Knee Arthroplasty | | | -3.5 [-15.8, 8.9] (p=0.583) | -4.5 [-12.3, 3.3] (p=0.257) |
| | End-Stage Hip Osteoarthritis | | | | -1.0 [-12.9, 10.8] (p=0.863) |

*Difference [95% confidence interval] in activity metrics adjusted for age, sex, body mass index, season, Charlson comorbidity index, education, ethnicity, alcohol consumption, and smoking status. A negative difference indicates lower activity in the cohort of interest relative to the reference cohort.*
